# Clinical, Lifestyle, Environmental and Dietary Determinants of Malnutrition in Adolescents on Antiretroviral Therapy in Ethiopia

**DOI:** 10.1101/2025.08.01.25332653

**Authors:** Meless Gebrie Bore, Lin Perry, Xiaoyue Xu, Andargachew Kassa Biratu, Marilyn Cruickshank

**Affiliations:** College of Medicine and Health Science, Faculty of Health, Hawassa University, Hawassa, Ethiopia; Faculty of Health, University of Technology Sydney, Ultimo, NSW, Australia; Prince of Wales Hospital. Randwick, NSW, Australia; School of Population Health, Faculty of Medicine, University of New South Wales, NSW, Australia; Sydney Children’s Hospital Network

**Keywords:** Nutritional status, Adolescents, Anti-Retroviral Therapy, HIV, Mixed methods study, Ethiopia

## Abstract

**Background:** Adolescence is a crucial period for growth, and adequate nutrition is essential for those living with HIV/AIDS and on antiretroviral therapy (ART). Despite declining HIV prevalence in Ethiopia, adolescents continue to face challenges that negatively impact their nutritional status. This study explored clinical, lifestyle, environmental, and dietary factors influencing the nutrition of adolescents on ART in selected Ethiopian hospitals.

**Method:** Cross-sectional surveys, clinical assessments and clinical record reviews were conducted, recruiting 384 ALHIV in receipt of ART at ten public hospitals in Addis Ababa and Oromia regions in August – December 2023. Participants were selected using proportionate random sampling for ALHIV and data were collected using a pre-tested interviewer-administered structured questionnaire and standardised assessments by trained healthcare workers.

**Results:** Nutritional assessments revealed 24.2% of participants classified as thin, 21.7% as stunted, and 34.9% as acutely malnourished. Factors significantly determinant of malnutrition included, for thinness, male gender, household food insecurity, a history of chronic infections such as tuberculosis, and symptom levels indicative of anxiety and moderate/moderately severe depression. Significant factors for acute malnutrition included younger rather than older adolescence (aged 10–17), male gender, larger household size (four or more members), household food insecurity, delayed disclosure of HIV status, history of chronic infections, generalized anxiety disorder, and low haemoglobin levels (<11 mg/dL).

**Conclusion and Recommendations:** Findings highlight the multifaceted challenges faced by ALHIV, inform and underscore the need for targeted nutritional and mental health interventions to address the specific challenges faced by this vulnerable group.

## 1. Introduction

Adolescence is a period marked by rapid physical, emotional and social development, making adequate nutritional status essential for this age group [1]. For adolescents living with HIV/AIDS, maintaining adequate nutrition is vital not only for overall health but also for the effectiveness of antiretroviral therapy (ART) [2]. Globally, HIV/AIDS remains a significant public health challenge and particularly among adolescents, who are uniquely vulnerable due to a combination of developmental, social, and economic factors. In Ethiopia, despite a decline in overall HIV prevalence—from 2.3% in 2002 to 0.8% in 2021 [3]—adolescents disproportionately bear the burden of the disease. This vulnerability is compounded by limited access to healthcare, pervasive stigma and engagement in high-risk behaviours [4].

Adolescents have distinct nutritional needs, and require increased energy, protein, vitamins, and minerals to support their growth and development [5]. However, many adolescents on ART face unique challenges, such as stigma, dietary restrictions and comorbidities, which can adversely affect their nutritional intake [6]. Additionally, socioeconomic factors, cultural practices, and difficulties accessing healthcare services play critical roles in determining the nutritional health of these individuals [7]. Inadequate nutritional status among these adolescents presents a critical barrier to achieving optimal health outcomes, impairing immune function and diminishing the efficacy of ART, leading to increased morbidity and mortality [8, 9].

Despite the Ethiopian Ministry of Health’s efforts to develop and implement adolescent and youth health programs, significant gaps persist [10]. While studies have emphasized the need for tailored nutritional interventions for individuals on ART [11], there remains a notable lack of comprehensive research focused specifically on adolescents in developing countries, including Ethiopia. This study aimed to identify the clinical, lifestyle, environmental and dietary factors associated with the nutritional status of adolescents on ART in selected hospitals in Ethiopia. Findings will contribute to the current body of knowledge to inform policymakers and healthcare providers about the specific nutritional needs and challenges faced by adolescents on ART. Addressing these factors will be essential to optimize the effectiveness of ART, enhance health outcomes, and promote the overall well-being of adolescents living with HIV in Ethiopia.

## 2. Methods

### 2.1 Study setting and period

This research was conducted in selected public health facilities providing ART for adolescents living with HIV (ALHIV) in Ethiopia. With the approval of the Human Research Ethics Committee at the University of Technology Sydney, Australia, (ETH23-7873), the Institutional Review Board of the College of Medicine and Health Sciences of Hawassa University (IRB/321/15), the Regional Health Bureau Ethics Review Committee, and the selected hospitals’ Ethics Review Committees in Ethiopia, the study took place in the Addis Ababa and Oromia regions from August to December 2023. A total of ten hospitals (i.e., Adama Referral Hospital, ALERT General Hospital, Asella Referral and Teaching Hospital, Batu Hospital, Bishoftu Hospital, Ras Desta Damtew Hospital, Shashamene Comprehensive Specialized Hospital, St Paul’s Comprehensive Specialized Hospital, Yekatit 12 Hospital, Zewditu Hospital) were chosen based on the high HIV prevalence in their catchment populations and substantial number of ART recipients available for recruitment.

### 2.2 Study design

The study employed an institutional-based mixed-methods design. The quantitative component primarily utilized survey design, supplemented by clinical assessments and the extraction of data from health records. Additionally, a small qualitative component used open-ended questions to provide deeper insights. Integrating quantitative and qualitative data, this research provides a holistic picture of the associated factors, barriers and facilitators to optimal nutrition in this vulnerable population.

To ensure methodological rigor and transparency in reporting, the paper adheres to the Strengthening the Reporting of Observational Studies in Epidemiology-Nutritional Epidemiology (STROBE-Nut) guidelines [12–14]. Operational definitions are provided in Supplementary file 1, Box 1).

### 2.3 Selection criteria and the sampling process

#### 2.3.1 Inclusion and exclusion criteria

The study included ALHIV aged 10–19 years on ART and attending ART units in ten selected hospitals across the Addis Ababa and Oromia regions of Ethiopia.

The study excluded adolescents who had only recently become aware of their HIV status and had been on ART for less than three months, as they may not have stabilized on treatment. Individuals with cognitive or communication deficits were also excluded to ensure accurate data collection and reporting. Adolescents under the age of 18 without parental or guardian consent were excluded to comply with ethical and legal standards. Furthermore, those without a medical registration number or unique ART number, or who could not provide their name and date of birth, were excluded due to the difficulties this posed for data reconciliation and verification.

A proportionate random sampling approach was employed to select study participants. To determine the sampling process for the target population, the total number of ALHIV receiving ART was sourced from the ART data clerk at each participating hospital. A sampling frame was constructed using the ART registration codes, with each participant assigned a unique research identifier to ensure confidentiality. Sample sizes were proportionally allocated according to the distribution of the population across the ten selected hospitals. Within each facility, eligible participants were randomly chosen by assigning a distinct registration number to each individual in the hospital’s records. The selection of participants was conducted using statistical software (SPSS version 26), which facilitated random selection based on these registration numbers.

#### 2.3.2 Sample size determination

The sample size was calculated based on nutritional status, factors shown to influence it, and food consumption patterns. The largest required sample size was chosen to ensure adequate power for all analyses, maintaining methodological rigor.

Sample size for ALHIV was calculated using a single population proportion formula 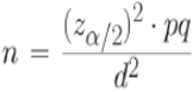 with a 5% marginal error (d) and CI of 95% (Z α/2= 1.96) based on the estimated proportion with undernutrition of 33.1% seen in 2020 in southern Ethiopia [15]. Based on these assumptions, the calculated sample size was 340.

For the factors influencing nutritional status, we applied a two-population proportion formula with a type one error of 5%, a power of 80%, and a 1:1 ratio of exposed to non-exposed groups, based on Sewale et al. (2018) and Shiferaw and Gebremedhin (2020). Using Epi Info version 7 software, the sample size for each variable was calculated, with the largest sample size of 352 selected for the variable “meal skipping.”

For evaluating food consumption and dietary patterns, the same formula was applied, using an estimated proportion of 50% due to a lack of prior studies, yielding a sample size of 384, Thus, the final sample size for assessing the nutritional status of ALHIV was set at 384.

### 2.4 Data collection

Data were collected during interviews using a pre-tested, interviewer-administered structured questionnaire for the target population. The questionnaire collected extensive quantitative data including socio-demographic details to identify factors influencing the nutritional status of ALHIV receiving ART. This dataset was augmented by clinical assessments and health record /clinical data extracted via health record/patient chart reviews, providing objective measurements related to health and nutrition.

Healthcare professionals, including nurses, health officers, public health practitioners, and nutritionists, conducted the interviews using smartphones and computers, with data entered using the Kobo Toolbox program. Before data collection, a three-day training session was held covering the study’s objectives, interview techniques, and methods for assessing nutrition including anthropometric measurements, clinical assessments and dietary evaluations.

The research team supervised the data collection process to ensure adherence to protocol. In case of technical issues with the platform, printed questionnaires were available. Daily checking of data completeness and accuracy, and data cleaning ensured quality control and data integrity. Consistency among raters was assessed using Cohen’s Kappa coefficient, with scores of 0.71 or higher indicating substantial agreement [27].

#### 2.4.1 Components of the questionnaire

The structured questionnaire was constructed using existing tools and literature, modified to align with the study objectives and context. It included both closed-ended and open-ended questions, which addressed sociodemographic characteristics, anthropometric, clinical (HIV and mental health), lifestyle, environmental, and dietary factors associated with the nutritional status of adolescents on ART.

Anthropometric measurements were collected as indicators of nutritional status. These measurements, along with clinical assessments, were conducted following standardized protocols to ensure consistency and accuracy. Weight was measured with digital scales, and height was recorded using portable stadiometers to calculate BMI. Circumferences (upper arm, waist, and hip) were assessed using a Holtain Measuring Tape, and skinfold thickness was measured at standardized sites (i.e., triceps, biceps, subscapular, and supra iliac) with Harpenden callipers to estimate body fat percentage. Body composition was evaluated using a handheld bioelectrical impedance analyser (BIA; Tanita BC-558) [16, 17]. Hand grip strength was assessed with a dynamometer (Jamar Hydraulic Hand Dynamometer, Model 5030J1) [18], recording the highest value from three squeezes of both hands. Reliability testing of anthropometric measurements was conducted using the Intraclass Correlation Coefficient (ICC), yielding a value of 0.856, which demonstrated good agreement among measurements.

Physical assessments were performed to identify common signs of malnutrition, such as wasting, stunting, bilateral pitting oedema, recent weight loss, dermatosis, palmar pallor, and specific eye signs like Bitot spots, corneal cloudiness, and corneal ulceration. Additionally, pallor in the palms, mucous membranes, and nail beds was evaluated. Wasting and skin-fold thickness were assessed using calibrated instruments to ensure precision [16].

Clinical HIV-related data retrieved manually from patient charts included clinical findings, laboratory results (haematological and biochemical markers), HIV status, CD4 levels, viral load, and nutritional deficiency-related issues. The WHO and Centres for Disease Control and Prevention (CDC) [19, 20] regard viral load assessment as a critical component of HIV/AIDS management as it can be used to improve treatment adherence, immune function, and nutritional status, ultimately leading to better health outcomes in this population. Routine assessment of CD_4_ levels and viral load at ART initiation provided insight into the overall health status and nutritional well-being of individuals living with HIV/AIDS [21]. Details of other health problems such as infections and co-morbidities were extracted from the health record.

Anxiety and depression were assessed using the validated Generalized Anxiety Disorder Questionnaire (GAD-7; Cronbach’s α = 0.77) and Patient Health Questionnaire-9 (PHQ-9; Cronbach’s α = 0.85) [22, 23].

Alcohol consumption was assessed as a lifestyle factor using the AUDIT-C screening tool [24, 25], which demonstrated high reliability with a Cronbach’s α of 0.847 in this study, consistent with previous research [26].

Open-ended questions drafted by the research team explored personal experiences and contextual factors that shape food choices and eating practices among ALHIV.

2.4.2 Questionnaire rigour The questionnaire was originally written in English and translated into Amharic; an independent language expert then back-translated the Amharic version into English, and versions were compared [28]. Local ART unit staff, including senior clinicians not employed at the study sites, reviewed both versions for content validity and relevance, and assessed the appropriateness of language for the intended audience. The Level Content Validity Index (I-CVI) and the Scale-Level Content Validity Index (S-CVI) were calculated [29, 30]. A threshold of S-CVI above 0.78 was set for good content relevance [31]. Six experts—two from academia, two from clinical practice, and two from a regional health office—rated item relevance and clarity on a four-point scale. The mean S-CVI was 0.981, indicating robust/strong initial content validity, with 98.1% of experts rating the items as moderately or highly relevant/clear. The experts also provided qualitative feedback, which was used to refine the clarity of the questionnaire.

### 2.5 Data processing and analysis

All statistical analyses were performed using SPSS version 24. Descriptive statistics were used to summarize the sociodemographic, clinical, lifestyle, environmental and dietary characteristics of these adolescents on ART. Frequency tables and figures were created to illustrate the distribution of each variable, ensuring accuracy and identifying any outliers or inconsistencies. Continuous variables were assessed for normality using the Kolmogorov-Smirnov and Shapiro-Wilk tests, with a significance level set at p <0.05, further validated through Q-Q plots. Missing values were assessed and excluded from the analysis, with only complete cases included to maintain data integrity.

Logistic regression analysis was conducted to identify determinants of nutritional status among ALHIV on ART. Variables with a p-value less than 0.25 in the univariate analysis were included in the multivariate logistic regression models using the backward elimination method, with statistical significance set at p < 0.05. All assumptions of the models were checked and met.

Qualitative data were coded into categorical variables and integrated with the quantitative dataset. These insights were analyzed alongside the quantitative data to provide a comprehensive picture of nutrition-related practices provided to adolescents by healthcare professionals, identified by study identifier numbers to preserve confidentiality.

## 3. Results

### 3.1 Participants’ characteristics

The study enrolled 384 ALHIV who were attending ART follow-up at 10 hospitals in Addis Ababa and Oromia regions. The response rate among eligible ALHIV invited to participate was 100%.

#### 3.1.1 Socio-demographic characteristics

Of all participants, 59.1% (n=227) were aged between 14 and 17 years, representing the middle age-range of adolescence, with a mean age of 15.9 years (± 2.19). More than half of the participants were female (54%, n=207). The majority (73%, n=281) were Orthodox Christians. Over two-thirds (66.1%, n=254) were recruited from Addis Ababa Regional Hospitals. Nearly all participants (98.7%, n=379) were students, with 46.6% (n=179) attending grades 1–8. In terms of household income, the largest proportion (45.6%, n=175) reported earning between 1000 and 3000 EBR monthly. More than half of the participants (51.6%, n=198) lived in households with 4–5 members (Table 1).

**Table 1.**
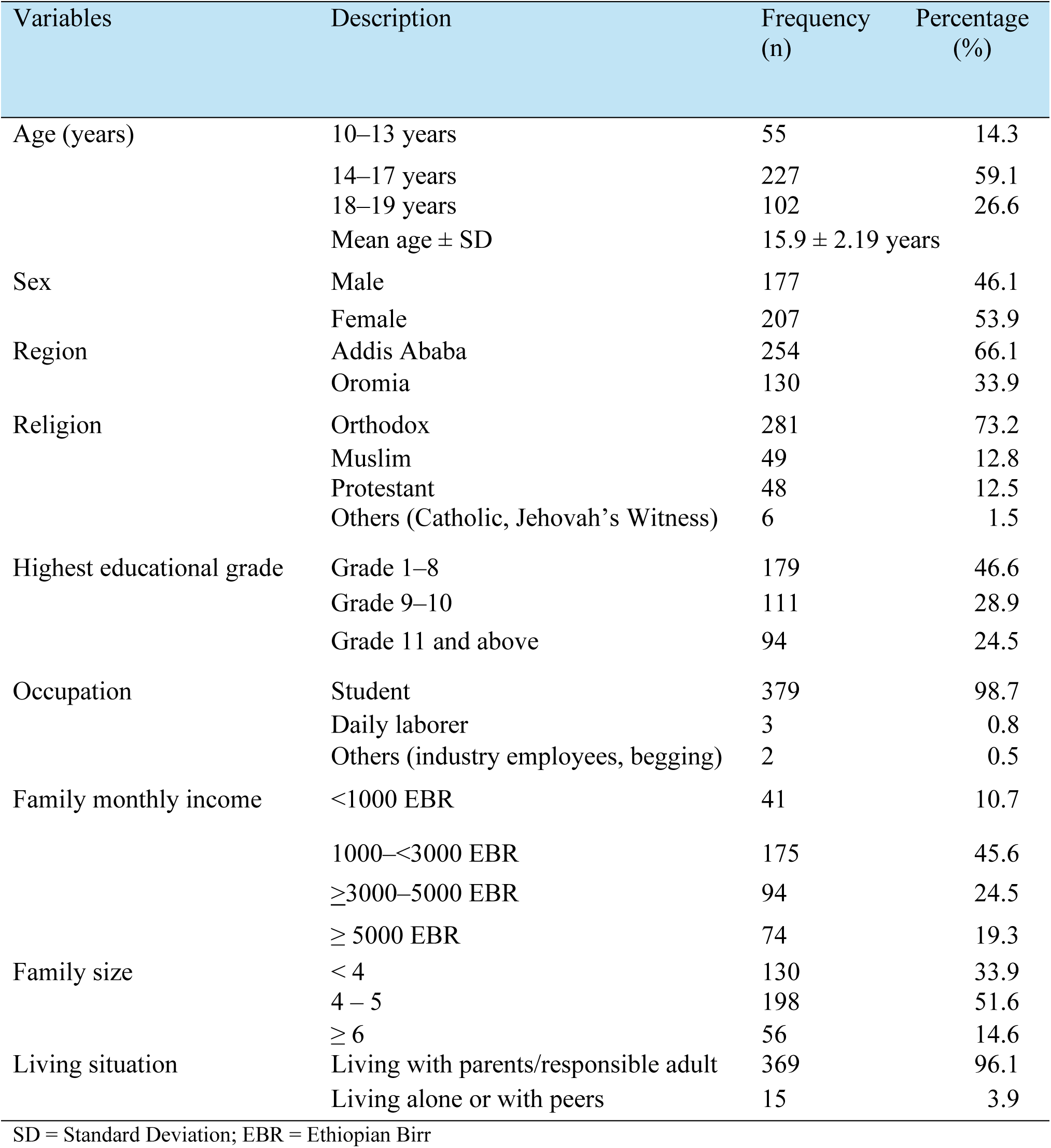
Socio-demographic characteristics of ALHIV attending ART follow-up in Ethiopia, 2024 (n=384)

#### 3.1.2 Anthropometric measures of nutritional status

The nutritional status of participants was assessed using three measures: Body Mass Index-for-Age Z-scores (BAZ), Height-for-Age Z-scores (HFA), and Mid-Upper Arm Circumference (MUAC) (Supplementary File, Box 1).

Regarding thinness, 24.2% (n=93) of participants had BMI-for-age Z-scores below −2 SD, with 26.9% (n=25) classified as severely thin (Z-scores < −3 SD) and 73.1% (n=68) as moderately thin (Z-scores between −3 SD and −2 SD), according to the WHO Growth Reference 2007 [32–34] (Table 2).

**Table 2:**
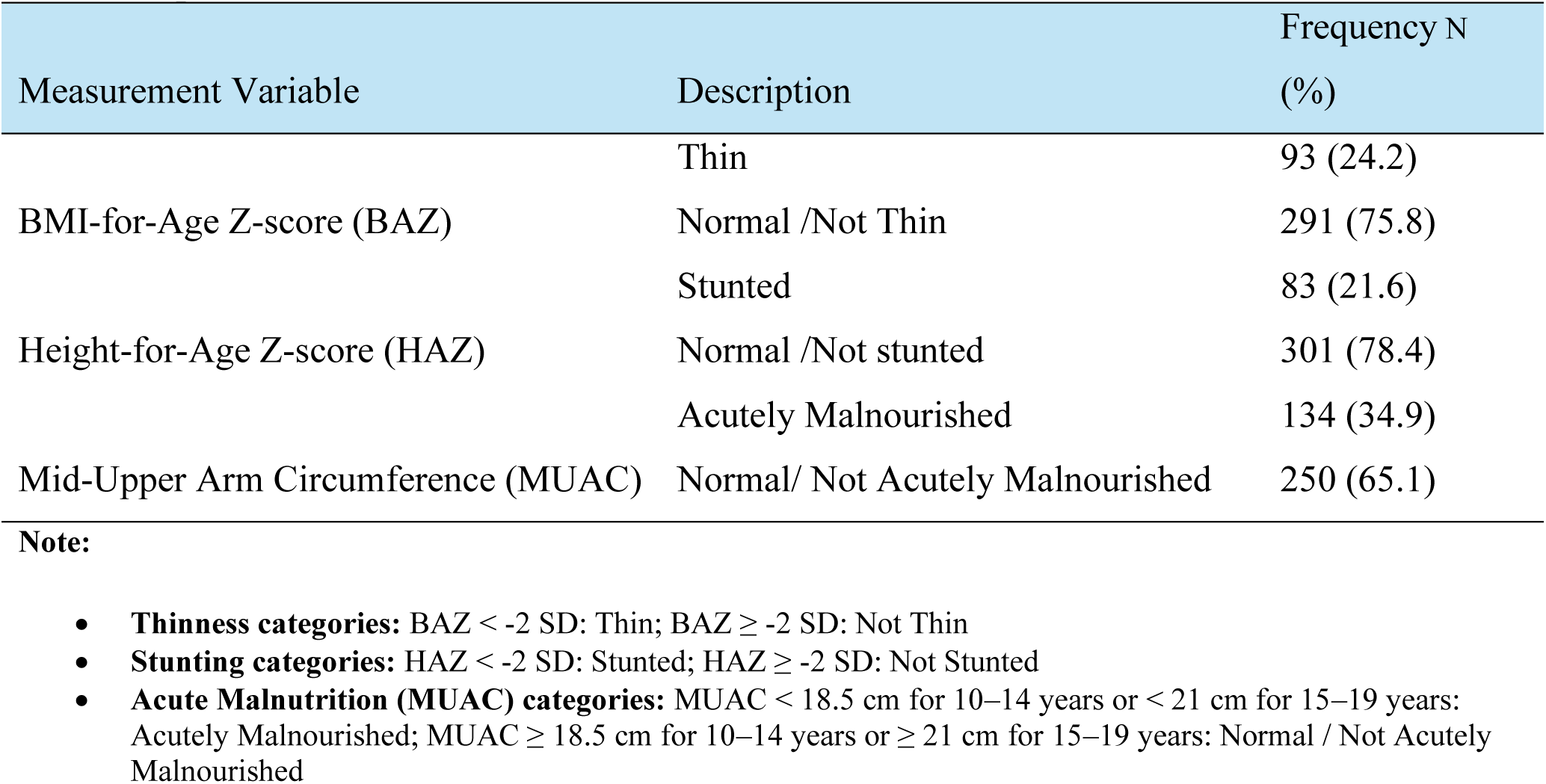
Nutritional status of adolescents living with HIV on ART follow-up in Ethiopia based on anthropometric measurements (n=384)

For stunting, 21.7% (n=83) of participants exhibited stunted growth (HFA Z-scores < −2 SD), with 28.9% (n=24) classified as severely stunted (Z-scores < −3 SD) [35] (Table 2).

For acute malnutrition, 34.9% (n=134) of participants were identified as malnourished based on MUAC measurements [36, 37]. Among this group, 80.6% (n=108) had moderate acute malnutrition, while 19.4% (n=26) were classified as having severe acute malnutrition (Table 2).

#### 3.1.3 Clinical characteristics HIV-related information

Of the 384 ALHIV participating in the study, 50.5% (n=194) had been diagnosed and commenced ART at the (median) age of 3.95 years/mean (SD) age 4.66 ± 3.7 years. Most (63%, n=242) had started ART 15 years ago or more. Overall, 79.9% (n=307) of the study participants had known their HIV status for four or more years; however, 47.4% (n=182) had a history of missed ART clinic appointments (Supplementary Table 1).

A significant majority (81.8%, n=314) of the study participants had HIV-positive family members in their households. Among these, 52.9% and 8.3% (n=166 and 26) reported that only one parent (mother or father, respectively) was HIV-positive, while 24.6% (n=76) had both parents living with HIV. For a few participants (13.9%, n=43) the whole nuclear family (e.g., both parents and sibling/s) were HIV-positive.

Almost all (98.7%, n=379) study participants had undergone viral load assessment within the previous three months. Of these participants, 97.7% had a viral load test result of less than 150 copies of viral load per ml of blood, and only 1% of the participants had 150 copies of viral load or more per ml of blood. Half of the participants had a CD_4_ level of 500–1500 cells/mm^3^ on ART enrolment and 36.7% (n=141) had a CD_4_ level below 500 cells/mm^3^ at the time of data collection.

Before starting ART, more than half of the participants were in clinical Stage I (n=247; 64.3%) and 29.4% (n=113) were in Stage III. After ART initiation, most participants (96.6%) were in clinical Stage I. This indicates a considerable improvement in their clinical status due to ART treatment. Most study participants (n=305; 79.4%) were taking first-line highly active anti-retroviral therapy (HAART) (Supplementary Table 2).

##### Mental health

Anxiety levels were assessed using the GAD-7 scale, with nearly half of the participants (49.5%, n=190) reporting anxiety. A small subset (8.1%, n=41) had moderate to severe anxiety, while 41.4% (n=159) experienced mild anxiety. Female participants reported higher levels of anxiety (28.4%, n=109) compared to males (21.1%, n=81), though the difference was not statistically significant (Figure 1 (A)). Anxiety levels varied by age, with 36.4% of early-age adolescents (10– 13 years) experiencing mild to severe anxiety, 49.8% of mid-aged adolescents (14–17 years), and 55.9% of late-age adolescents (18–19 years) (Figure 1 (B)).

**Figure 1:**
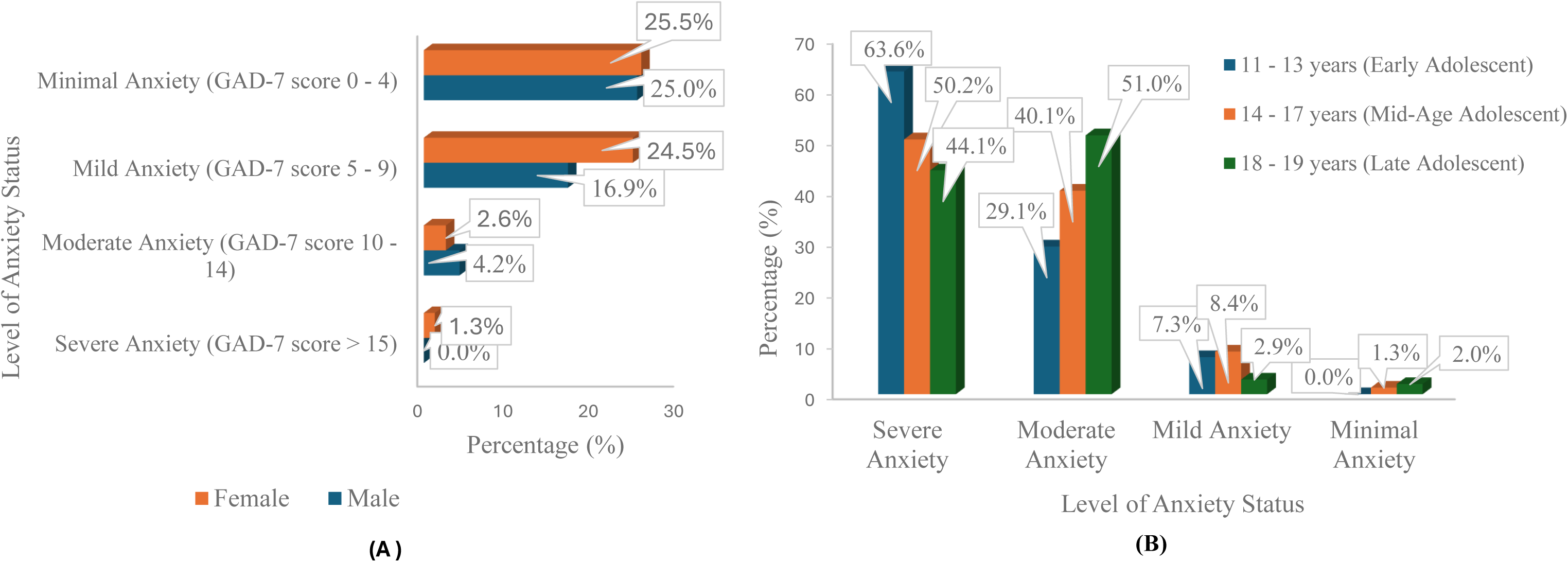
Prevalence of anxiety assessed using the Generalized Anxiety Disorder 7-item scale (GAD-7) by gender (A) and age category (B) among ALHIV on ART follow-up in Ethiopia 2024 (n=384)

Scores indicative of depression, measured using the PHQ-9 scale, were reported by 27.6% (n=106) of participants, with 6.8% (n=26) experiencing moderate to severe depression. A higher proportion of female participants reported depressive symptoms (27.6%, n=106) compared to males (19.8%, n=76) (Figure 2 (A)). Depression levels also varied by age group, with 47.3% of early-age adolescents (10–13 years), 46.7% of mid-aged adolescents (14–17 years), and 49.0% of late-age adolescents (18–19 years) reporting mild to moderate depressive symptoms (Figure 2 (B)).

**Figure 2:**
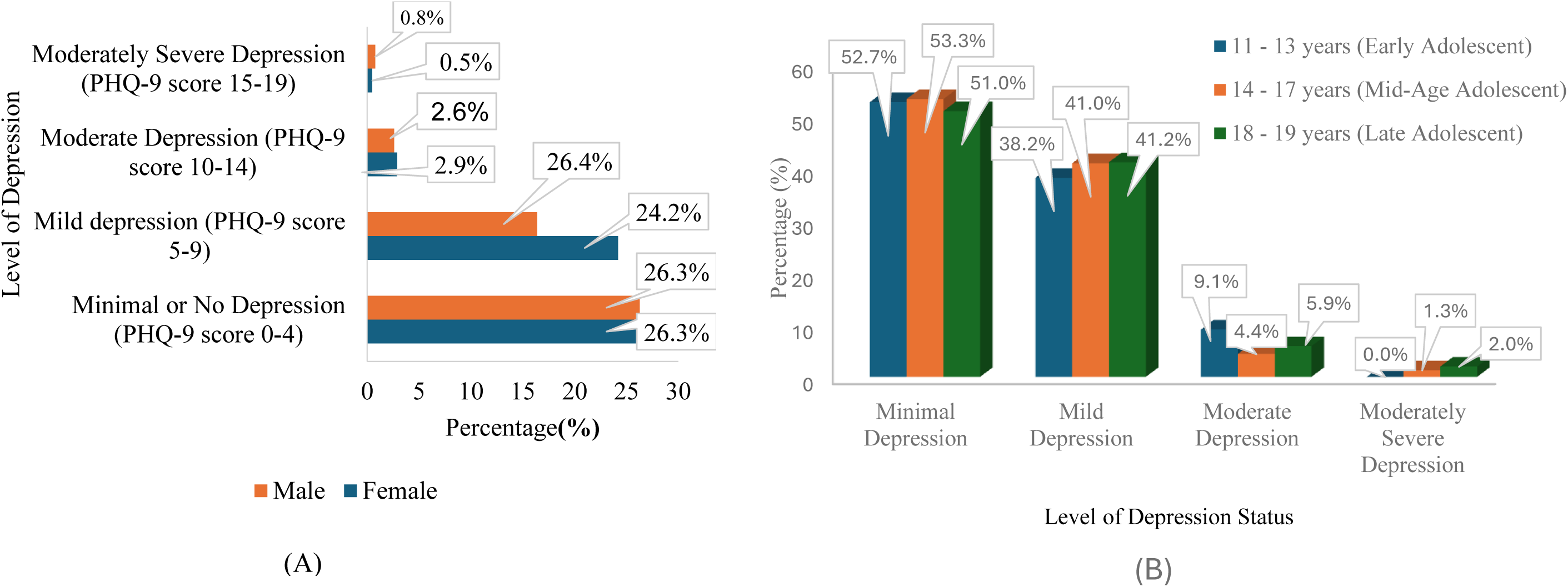
Distribution of depression assessed using a modified PHQ-9 by gender (A) and age category (B) among ALHIV on ART follow-up in Ethiopia 2024 (n=384)

##### Other health problems

Most participants (81.5%; n=313) had experienced opportunistic infections at some point in their lives. Of these, 57.5% (n=221) had experienced chronic respiratory illnesses such as colds, pneumonia, and tuberculosis. Notably, one third of these participants (33.1%, n=127) had a history of tuberculosis and were being treated with an anti-tuberculosis regimen. Other health problems were also common. Over half had experienced gastrointestinal system problems (56.5%, n=217) such as salmonellosis and gastritis/peptic ulcer disease, dermatologic problems (46.4%, n=178), and central nervous system problems (33.3%, n=128) such as headaches, meningitis, epilepsy and Guillain-Barré Syndrome. Furthermore, half of the participants had experienced some form of illness in the previous three months. Of these participants, 67.2% had experienced respiratory system problems, while 19.3% (n=74) had suffered gastrointestinal system problems (Supplementary Table 1).

#### 3.1.4 Lifestyle-related characteristics - alcohol use

Approximately one-third of participants (29.9%, n=115) had ever used alcohol, with 9.9% (n=38) reporting current alcohol consumption. Of these, 34.2% (n=13) of the 38 participants showed risky, high-risk, or addictive drinking patterns (Table 3). Males had higher rates of risky alcohol use (26.4%, n=10) compared to females (7.9%, n=3). Among age groups, 23.7% (n=9) of later-age adolescents (18–19 years) engaged in risky or addictive drinking, with 7.9% (n=3) classified as high-risk. In contrast, 10.5% (n=4) of mid-age adolescents (14–17 years) displayed risky drinking behaviours.

**Table 3.** Alcohol consumption risk category of ALHIV on ART follow-up in Ethiopia using the *AUDIT screening test, 2024 (n=38)

| Variables | Frequency N (%) |
| --- | --- |
| Low risk (AUDIT Score 1 – 7) | 25 (65.8) |
| Risky (AUDIT Score 8 – 15) | 10 (26.3) |
| High Risk (AUDIT Score 16 - 19) | 1 (2.6) |
| Addiction Likely (AUDIT Score 20 - 40) | 2 (5.3) |

\*AUDIT - Alcohol Use Disorders Identification Test

#### 3.1.5 Environmental issues and dietary considerations

Food security emerged as a significant concern among the participants. Approximately 34.4% (n=132) reported insufficient food availability in their households, and over half (53.1%, n=204) involuntarily ate a restricted variety of foods. Additionally, 54.2% (n=208) were unable to consume their preferred foods due to resource limitations, while 40.1% (n=154) reported eating foods they did not want and consuming smaller meals because of food scarcity. Nearly half (44.8%, n=172) had fewer meals per day than desired, and 25.8% (n=99) reported having periods with no food at home due to resource constraints. Furthermore, 55.3% (n=212) went to bed hungry at least 1–3 times per week, and 15.4% (n=59) experienced a full day and night without food. A large proportion (83.6%, n=321) indicated a history of meal-skipping, and more than half (53.1%, n=204) lived in food-insecure households (Supplementary Table 3).

Regarding food preservation, most participants employed household preservation techniques, including canning (91.9%, n=353) and drying (79.9%, n=307). Additionally, about half of the participants used refrigeration, freezing, and salting methods to preserve food.

Most participants (77.3%; n=297) had access to clean water at home. Among the 87 participants without this, nearly half (48.3%, n=42) treated their water to ensure it was safe for drinking and other uses. The primary methods of water treatment included the use of chlorine-based solutions and water filters. Regarding sanitation facilities, most participants (82%, n=315) had pit latrines in their households, while a smaller proportion (17.2%, n=66) had access to flush toilets. Only 0.3% (n=1) of participants practiced open-field defecation (Supplementary Table 4).

In addition to food insecurity, feeding-related health issues were common, with approximately 28.6% (n=110) of participants experiencing symptoms such as vomiting, diarrhea, and loss of appetite. Nutritional supplementation was provided to 62% (n=238) of participants, who received Ready-to-Use Therapeutic Food (RUTF) from their ART clinic, with more than half receiving one sachet of 92g RUTF per day for up to two months (Supplementary Table 1).

### 3.2 Determinant factors for malnutrition

#### 3.2.1 Determinants of thinness

A regression analysis was conducted to explore potential determinants of malnutrition defined as thinness (i.e. BMI-for-age Z-score values below −2 SD). Correlation analysis was conducted to determine the potential variables fitted for the model (Supplementary Table 5). From all the categories of variables tested, 10 variables met the inclusion criterion of p-value < 0.25 in univariate analysis. These variables included sociodemographic factors (age, sex/gender, household monthly income), clinical and HIV-related characteristics (history of nutritional assessment during ART initiation, duration of HIV awareness, history of long-lasting infections such as tuberculosis, GAD [anxiety] and PHQ-9 [depression]), and environmental and dietary factors (food insecurity concern, history of nutritional supplement use), in relation to thinness.

##### Sociodemographic variables

Among the sociodemographic variables analyzed, only sex showed a significant association with thinness. Female ALHIV were 73% less likely to be thin or malnourished compared to their male counterparts (AOR = 0.27; 95% CI: 0.16, 0.45). In contrast, age and family monthly income were not significantly associated with thinness (Table 4).

**Table 4.**
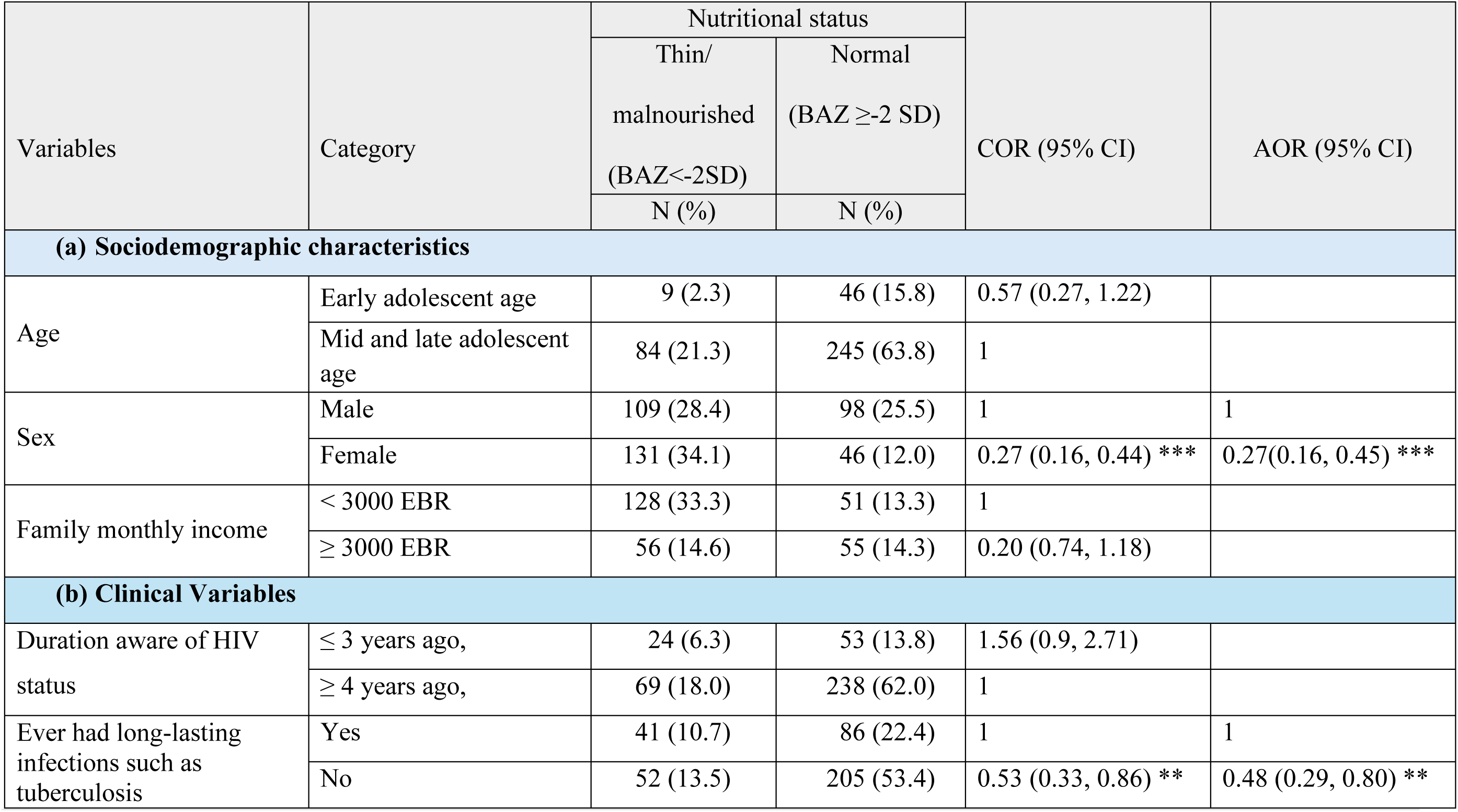
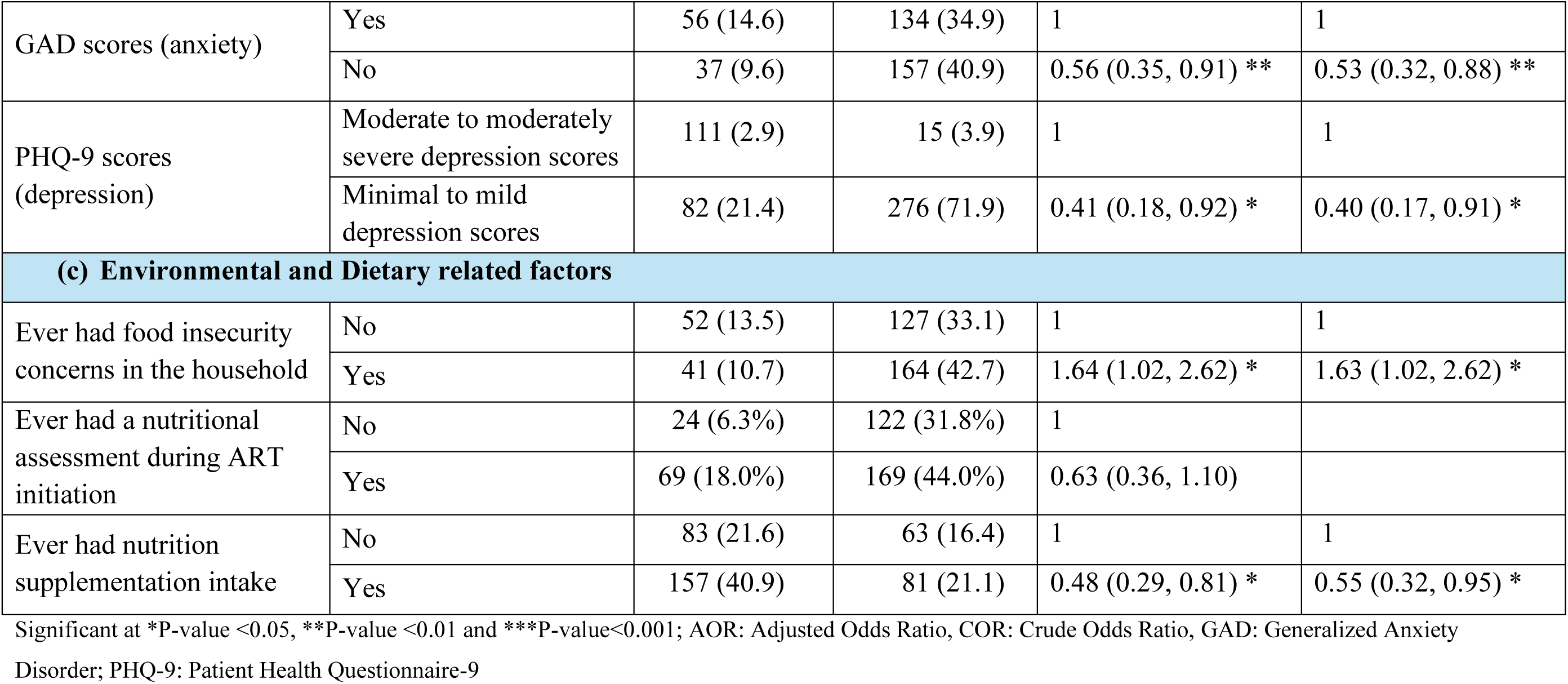
Factors significantly associated with thinness/malnutrition (BAZ) among ALHIV on ART in Ethiopia, 2024 (n=384)

##### Clinical variables

ALHIV who had never experienced chronic infections, such as tuberculosis, were 52% less likely to be thin or malnourished than those who had suffered from such infections (AOR = 0.48; 95% CI: 0.29, 0.80). This highlights the potential link between chronic health conditions and nutritional outcomes (Table 4).

ALHIV without symptoms of Generalized Anxiety Disorder (GAD), as indicated by GAD-7 scores below 5, were 47% less likely to be thin or malnourished compared to those with higher anxiety scores (AOR = 0.53; 95% CI: 0.32, 0.88). Additionally, ALHIV experiencing minimal to mild symptoms of depression (PHQ-9 scores < 9) were 60% less likely to be thin or malnourished than those with moderate to severe depression scores (AOR = 0.40; 95% CI: 0.17, 0.91). In contrast, factors such as history of nutritional assessment during ART initiation and duration of HIV awareness, were not significantly associated with thinness (Table 4).

##### Lifestyle factor

In this study, alcohol consumption did not exhibit a significant relationship with thinness.

##### Environmental and Dietary factors

Food security was significantly linked to thinness. Adolescents living with HIV who experienced food insecurity were 1.63 times more likely to be thin or malnourished than those who did not face such challenges (AOR = 1.63; 95% CI: 1.02, 2.62). Conversely, ALHIV who received nutritional supplementation had a 45% lower likelihood of being thin or malnourished compared to those who did not receive such support (AOR = 0.55; 95% CI: 0.32, 0.95) (Table 4). No other environmental and dietary-related factors were significantly associated with thinness.

#### 3.2.2 Determinants of acute malnutrition

A regression analysis was conducted to explore potential determinants of acute malnutrition, defined as MUAC < 18.5 cm for 10–14 years or < 21 cm for 15–19 years (Supplementary File Box 1). Correlation analysis was conducted to determine the potential variables for entry in the model (Supplementary Table 6). From all the categories of variables tested, 13 variables met the inclusion criterion of p-value < 0.25 in the univariate analysis. These variables included sociodemographic variables (age, sex, family size), clinical variables (anxiety status, haemoglobin, alanine aminotransferase level (SGPT), duration aware of HIV status, longstanding disease), and environmental and dietary factors (ever had food insecurity in household, number of meals eaten in 24 hours, ever had nutrition supplementation), which were significantly associated with acute malnutrition among ALHIV.

##### Sociodemographic variables

Early and mid-aged ALHIV were more likely to suffer from malnutrition than late-age ALHIV. Malnutrition risks were 1.94 times higher in ALHIV aged 10–17 years (AOR = 1.94; 95% CI: 1.03, 3.64). Female adolescents with HIV were 42% less prone to being malnourished than their male counterparts (AOR = 0.58; 95% CI: 0.38, 0.88). ALHIV residing in households with three or fewer family members were 56% less likely to experience malnutrition than those living in households with four or more members (AOR = 0.44; 95% CI: 0.24, 0.81) (Table 5).

**Table 5.** Factors associated with acute malnutrition using MUAC-for-age indices among ALHIV on ART in Ethiopia, 2024 (n=384)

| Variables | Category | Nutritional Status using MUAC-for-Age indices |  | COR (95% CI) | AOR (95% CI) |
| --- | --- | --- | --- | --- | --- |
|  |  | Acutely Malnourished | Not acutely Malnourished |  |  |
|  |  | N (%) | N (%) |  |  |
| (a) Sociodemographic Characteristics |  |  |  |  |  |
| Age | Early and mid-adolescent age (10–17 years) | 106 (27.6) | 176 (45.8) | 1.59 (0.97, 2.62) * | 1.94 (1.03, 3.64) * |
|  | Late adolescent age (18–19 years) | 28 (7.3) | 74 (19.3) | 1 | 1 |
| Sex | Female | 61 (15.9) | 146 (38.0) | 0.59 (0.39, 0.91) * | 0.58 (0.38, 0.88) * |
|  | Male | 73 (19.0) | 104 (27.1) | 1 | 1 |
| Household monthly income | < 3000 EBR | 82 (21.4) | 134 (34.9) | 1.36 (0.89, 2.09) |  |
|  | ≥ 3000 EBR | 52 (13.5) | 116 (30.2) | 1 |  |
| Family size | Less than or equal to three | 35 (9.1) | 95 (24.7) | 0.58 (0.36, 0.92) * | 0.44 (0.24, 0.81) * |
|  | Four and above | 99 (25.8) | 155 (40.4) | 1 | 1 |
| (b) Clinical Variables |  |  |  |  |  |
| Duration aware of HIV status | ≤ 3 years ago | 42 (10.9) | 35 (9.1) | 2.80 (1.68, 4.67) *** | 2.96 (1.62. 5.40) *** |
|  | ≥ 4 years ago | 92 (24.0) | 215 (56.0) | 1 | 1 |
| PHQ-9 scores (depression) | Minimal/no depression scores | 64 (16.7) | 138 (35.9) | 0.74 (0.49, 1.13) |  |
|  | Mild to severe depression scores | 70 (18.2) | 112 (29.2) | 1 |  |
| GAD scores (anxiety) | No | 58 (15.1) | 136 (35.4) | 0.64 (0.42, 0.98) * | 0.50 (0.28, 0.89) * |
|  | Yes | 76 (19.8) | 114 (29.7) | 1 | 1 |
| Ever had longstanding disease (e.g., tuberculosis) | No | 57 (14.8) | 70 (18.2) | 0.53 (0.34, 0.82) ** | 0.52 (0.33, 0.81) ** |
|  | Yes | 77 (20.1) | 180 (46.9) | 1 | 1 |
| Haemoglobin status | <11.5 gm/dl | 26 (7.2) | 29 (8.1) | 3.59 (1.06, 12.12) * | 7.29 (1.31, 40.5) * |
|  | 11.5–15.0 gm/dl | 98 (27.3) | 186 (51.8) | 1.70 (0.95, 3.05) | 1.92 (0.83, 4.42) |
|  | >15.0 gm/dl | 4 (1.1) | 16 (4.5) | 1 | 1 |
| Alanine transaminase (SGPT) | 0–50 U/L | 86 (36.8) | 135 (57.7) | 0.40 (0.13, 1.26) * | 0.29 (0.09, 0.99) * |
|  | >50 U/L | 8 (3.4) | 5 (2.1) | 1 | 1 |
| (c) Environmental and Dietary related factors |  |  |  |  |  |
| Ever had food insecurity concerns in households | Yes | 64 (16.7) | 141 (28.4) | 1.46 (0.96, 2.22) * | 1.85 (1.09, 2.14) * |
|  | No | 70 (18.2) | 109 (28.4) | 1 | 1 |
| Number of meals eaten in 24 hours (day and night) | ≤ 2 meals | 30 (7.8) | 30 (7.8) | 2.11 (1.21, 3.69) * | 1.86 (1.08, 3.19) * |
|  | 3–5 meals | 104 (27.1) | 220 (57.3) | 1 | 1 |
| Ever had nutrition supplementation intake | No | 39 (10.2) | 107 (27.9) | 0.55 (0.35, 0.86) ** | 0.45 (0.36, 0.79) ** |
|  | Yes | 95 (24.7) | 143 (37.2) | 1 | 1 |

Significant at \*P-value <0.05, \*\*P-value <0.01 and \*\*\*P-value<0.001; AOR: Adjusted Odds Ratio, COR: Crude Odds Ratio, GAD: Generalized Anxiety Disorder; PHQ-9: Patient Health Questionnaire-9

##### Clinical variables

ALHIV without anxiety symptoms (GAD-7 score < 5) were 50% less likely to develop malnutrition than those with above-threshold anxiety scores (GAD-score ≥5) (AOR = 0.50; 95% CI: 0.28, 0.89). ALHIV who had known their HIV status for three years or less (AOR = 2.96; 95% CI: 1.62, 5.40) had a 2.96 times higher risk of malnutrition than those who became aware four or more years earlier.

ALHIV with haemoglobin levels below 11.5 grams per decilitre were 7.29 times more likely to record malnutrition than those with haemoglobin levels exceeding 15 grams per decilitre (AOR = 7.29; 95% CI: 1.31, 40.5). ALHIV with SGPT levels between 0 and 50 U/L were 71% less likely to experience malnutrition (AOR = 0.29; 95% CI: 0.09, 0.99), than those with SGPT levels of 50 U/L or higher.

ALHIV who had never experienced a prolonged illness such as tuberculosis were 48% less likely to experience malnutrition than those who had (AOR = 0.52; 95% CI: 0.33, 0.81) (Table 5).

##### Environmental and Dietary-related factors

ALHIV who had experienced food insecurity within their households ran a 1.8-fold higher risk of malnutrition (AOR = 1.85; 95% CI: 1.09, 2.14) than those who had never encountered such issues. In addition, malnutrition was more prevalent among ALHIV who consumed fewer meals per day than required. The odds of malnutrition were 1.86 times higher in ALHIV who had ever eaten two meals or fewer in a day than in those who had always consumed three or more meals daily (AOR = 1.86; 95% CI: 1.08, 3.19). Furthermore, ALHIV receiving nutritional supplementation were 55% less likely to experience malnutrition than those without supplementation (AOR = 0.45; 95% CI: 0.36, 0.79) (Table5).

#### 3.2.3. Determinants of Stunting

Sociodemographic, clinical, lifestyle, and environmental and dietary factors were examined but none showed a statistically significant association with stunting in adolescents living with HIV.

## 4. Discussion

In adolescence, a crucial phase of growth and development, the body’s nutritional demands increase significantly, and it is essential that these are met for the overall health of this future adult population and the optimal management of HIV [38]. Study findings reveal the multifaceted nutritional and health challenges faced by ALHIV in Ethiopia. They demonstrate substantial rates of malnutrition, in relation to thinness (24.2%), stunting (21.7%), and acute malnutrition (34.9%), suggestive of widespread deficiencies that can impact physical and cognitive development as well as overall well-being. These findings underscore the public health concern surrounding malnutrition among ALHIV.

The prevalence of thinness observed in this study (24.2%) aligns with findings from studies from Uganda and southern Ethiopia [15, 39] but exceeds rates in Nigeria and more recent Ugandan studies [40, 41]. Similarly, the prevalence of stunting (21.7%) is comparable to recent studies in Uganda (25%) (Dave et al., 2022) but lower than those in southern Ethiopia (33.1%) and earlier Ugandan findings (36%) (Francis et al., 2015; Shiferaw & Gebremedhin, 2020). These discrepancies may reflect differences in socio-economic conditions, access to healthcare, and local dietary practices. While the lower rates of stunting in this study suggest relatively better nutritional support or healthcare access, they still highlight a significant public health challenge and underscore how wide-spread this is. Addressing the underlying socio-economic determinants will be crucial to improving nutritional outcomes and tackling malnutrition in this vulnerable population.

This study also emphasizes the complexity of malnutrition among ALHIV, highlighting age, gender, and family size influencing nutritional outcomes. Younger adolescents (aged 10–17 years) were more likely to experience acute malnutrition, likely due to the increased nutritional demands of growth during this pubertal developmental period [34, 42]. The rapid physical and cognitive development in this age group amplifies their nutritional needs, which can be further exacerbated by HIV-related illness and ART side effects [43]. These findings underscore the importance of developing age-targeted interventions to address the specific nutritional needs of early and mid-age adolescents to prevent long-term developmental deficits. Given that most of those affected were school students, school health programs may provide a route to implement such interventions [44].

Gender differences in nutritional status were also observed, with male ALHIV more vulnerable to malnutrition than females. This may be due to higher caloric needs from rapid muscle growth, poorer appetite, or ART-related side effects. In contrast, female ALHIV were less vulnerable, possibly due to lower caloric requirements or better access to healthcare and nutrition. These findings emphasize the need for gender-specific intervention**s** to address the unique nutritional challenges faced by male ALHIV, ensuring tailored support to improve nutritional status and long-term health outcomes.

The role of family size in nutritional status suggests that smaller households may be better able to provide adequate resources, including food, to meet the nutritional needs of ALHIV. This finding underscores the broader social determinants of health, such as household structure and socio-economic status, in influencing the nutritional status of vulnerable populations. While family income was not directly linked to malnutrition, the fact that smaller families were less likely to experience malnutrition suggests that household resource distribution may be an important factor in the nutritional health of ALHIV. Interventions that consider the socio-economic context, including household size and resource availability, could improve the effectiveness of nutritional support for ALHIV.

Chronic infections and health markers were also crucial factors in the nutritional status of ALHIV. The absence of chronic infections such as tuberculosis was associated with a lower likelihood of malnutrition, which is consistent with other research showing that infections contribute to poor nutritional outcomes by increasing energy expenditure and impairing nutrient absorption [45, 46]. Additionally, haemoglobin levels were found to be a strong predictor of malnutrition, with lower levels significantly increasing the likelihood of nutritional deficiencies. This aligns with studies indicating that anaemia, often caused by HIV or nutritional deficiencies, is associated with higher rates of malnutrition [47]. Similarly, liver function, as indicated by SGPT levels, was found to relate to risk of malnutrition, with elevated SGPT levels suggesting liver damage that may accompany increased risk, while normal SGPT levels, reflecting better liver function, may indicate a lower likelihood of malnutrition. This finding flags a potential vicious circle, with liver health compromised by malnutrition, but crucial in maintaining nutritional balance as liver dysfunction can lead to metabolic disturbances that exacerbate malnutrition [48]. These findings highlight the need for comprehensive healthcare management for ALHIV, including regular monitoring and intervention for infections, anaemia, and liver dysfunction, to improve nutritional status and overall health.

Food insecurity emerged as a significant environmental issue, with participants reporting low food variety and meal frequency, which exacerbated their malnutrition risk. Those experiencing food insecurity were more likely to be thin, reflecting previous research that links food insecurity to poor nutritional outcomes among HIV-positive individuals [15, 49]. Conversely, nutritional supplementation was linked to significantly lower rates of malnutrition among ALHIV, highlighting the importance of targeted nutritional programs as part of comprehensive HIV care. Dietary patterns also played a vital role, with participants who consumed three or more meals per day less likely to be malnourished. This finding underscores the importance of promoting healthy eating habits, improving access to nutritious foods, and encouraging consistent meal consumption to combat malnutrition [50], supported by targeted nutritional programs.

Environmental factors such as inadequate access to clean water and poor sanitation further complicate the nutritional landscape. The association between poor sanitation and increased infection risk suggests that improving water and sanitation infrastructure is essential for promoting better health outcomes among ALHIV [51, 52]. Furthermore, clinical indicators such as elevated SGPT levels and the presence of chronic illnesses like tuberculosis also correlated with malnutrition, emphasizing the importance of integration of clinical and nutritional management strategies [53, 54].

Interestingly, no factors were found to be significantly associated with stunting (HFA indices), indicating an important area for further research. This suggests that underlying or long-term factors not captured by this study may contribute to stunting in ALHIV receiving ART. Potential factors could include early childhood malnutrition, genetic predispositions, or the prolonged effects of ART on growth. Future research should explore chronic malnutrition, early growth patterns, and the long-term impacts of ART on stunting in this population to better understand the mechanisms involved.

In summary, the findings suggest that food insecurity, poor dietary diversity, inadequate healthcare, and socio-economic factors are significantly affecting the nutritional outcomes of ALHIV, including the shockingly high prevalence of malnutrition revealed by this study. These underlying issues may exacerbate the nutritional deficiencies observed among ALHIV, who face compounded health challenges due to their HIV status. Addressing these disparities requires a holistic approach that integrates improved nutrition, enhanced healthcare access, and targeted support for ALHIV. Given that nearly all participants in this study were students, integrating HIV management including nutritional care into school health programs may offer an effective strategy to improve adolescent health outcomes, as demonstrated in other adolescent health interventions [55–57].

### 4.1 Limitations

This study encountered several limitations that may influence the interpretation of its findings. A major concern was the reliance on self-reported data from ALHIV which could introduce response or social desirability bias and recall bias could influence participants’ accuracy in recounting past experiences. While efforts were made to minimize these biases through triangulation and standardized protocols, their potential impact on data accuracy remains a concern. Future research could enhance reliability by incorporating objective measures and cross-validation techniques.

Excluding family/ caregiver insights may have overlooked crucial support dynamics in the lives of ALHIV. Caregivers play a crucial role in managing nutrition and health, and their perspectives on food security and healthcare access could provide a more complete understanding of the factors influencing nutritional outcomes. Future studies should include caregiver input for a more holistic view of ALHIV’s support systems.

The cross-sectional design limited the ability to establish causal relationships between nutritional practices and health outcomes; longitudinal or interventional studies could offer further insights into these dynamics.

Lastly, socio-political and economic challenges within the study context, along with the disruptions caused by the COVID-19 pandemic, may have affected the results. The long-term effectiveness of healthcare programs has also not been evaluated. Addressing these limitations through larger sample sizes, longitudinal approaches, and improved data collection methods, while considering the ongoing impact of the pandemic, could lead to more robust findings regarding the impact of nutritional practices on ALHIV.

## 5. Conclusion and recommendations

This study underscores the complex nutritional challenges faced by ALHIV in Ethiopia, highlighting significant rates of malnutrition, including thinness, stunting, and acute malnutrition. These challenges are influenced by socio-demographic factors, food insecurity, chronic infections, and mental health issues. Revealing the scope and scale of underpinning influences, the study emphasizes the urgent need for integrated nutritional support and healthcare interventions that address these interrelated factors.

The findings reinforce the importance of age- and gender-specific interventions, particularly for younger adolescents and male ALHIV, who face higher malnutrition risks. Further research should be conducted to determine whether tailoring programs to meet the unique needs of these groups will improve long-term health outcomes. Additionally, addressing food insecurity, poor dietary diversity, and promoting consistent meal consumption patterns are critical for improving nutritional status.

The role of chronic infections, anaemia, and liver dysfunction in malnutrition highlights the necessity for comprehensive healthcare management that includes regular monitoring and treatment of these factors. Furthermore, improving sanitation and access to clean water will reduce the environmental factors that exacerbate malnutrition risks.

To improve ALHIV’s health and nutritional status, healthcare programs should integrate nutritional support, mental health services, and treatment for chronic infections. Targeted interventions should focus on age and gender differences, and healthcare workers must receive training to provide effective care. Future research should adopt longitudinal studies to better understand the long-term effects of nutritional practices on ALHIV health.

Lastly, the integration of HIV management within school health programs could enhance access to health resources for adolescents, as seen in other global adolescent health initiatives. Addressing socio-economic disparities through policy advocacy will also be key in ensuring that the nutritional and health needs of ALHIV are met.

## Data Availability

The data supporting the findings of this study are available upon reasonable request. Access to the data may be restricted to ensure participant confidentiality and to comply with ethical guidelines.

## Declaration

### Ethics approval

This study was conducted in accordance with the Declaration of Helsinki. Ethical approval was obtained from the Human Research Ethics Committee at the University of Technology Sydney, Australia (Ref. Number: ETH23-7873); the Institutional Review Board of the College of Medicine and Health Sciences, Hawassa University, Ethiopia (Ref. Number: IRB/321/15) — which also covered the Oromia region study sites; the Addis Ababa City Administration Health Bureau Ethics Review Committee, Ethiopia (Ref No: A/A/3/54/227); and St. Paul’s Hospital Millennium Medical College Institutional Review Board (Ref No: PM23/181). Permission was secured from all health institutions involved in the study. Written informed consent for participation was obtained from participants’ legal guardians or next of kin. Confidentiality was assured and maintained throughout the study.”

### Conflict of Interest

The authors declare that there is no conflict of interest.

### Funding

The authors received no specific funding for this work. It is part of a PhD thesis by MGB. MGB is a higher degree research candidate at UTS, supported by the International Research Training Program (IRTP). The IRTP is a commonwealth scholarship funded by the Australian government and the Department of Education and Training.

### Authors contribution

MGB performed data collection, analysis and drafted the initial manuscript. All authors contributed to the conceptualisation, methodology development, data analysis, interpretation, revision and approval of the final manuscript.

## Acknowledgments

The authors wish to acknowledge the Local Government Areas of Ethiopia, the Health Bureau, and all hospital administrators for granting permission to collect data. We also extend our gratitude to the ART unit staff members and patients for their participation in the study.

## Notes

### Competing Interest Statement

The authors have declared no competing interest.

### Funding Statement

The authors received no specific funding for this work. It is part of a PhD thesis by MGB. MGB was a higher degree research candidate at UTS, supported by the International Research Training Program (IRTP). The IRTP is a commonwealth scholarship funded by the Australian government and the Department of Education and Training.

### Author Declarations

This study was conducted in accordance with the Declaration of Helsinki. Ethical approval was obtained from the Human Research Ethics Committee at the University of Technology Sydney, Australia (Ref. Number: ETH23-7873) the Institutional Review Board of the College of Medicine and Health Sciences, Hawassa University, Ethiopia (Ref. Number: IRB/321/15) — which also covered the Oromia region study sites the Addis Ababa City Administration Health Bureau Ethics Review Committee, Ethiopia (Ref No: A/A/3/54/227) and St. Paul’s Hospital Millennium Medical College Institutional Review Board (Ref No: PM23/181). Permission was secured from all health institutions involved in the study. Written informed consent for participation was obtained from participants’ legal guardians or next of kin. Confidentiality was assured and maintained throughout the study.

## References

[1] Patton GC, Sawyer SM, Santelli JS, Ross DA, Afifi R, Allen NB, et al. Our future: a Lancet commission on adolescent health and wellbeing. Lancet. 2016;387(10036):2423–78.

[2] WHO. HIV and adolescents: HIV testing and counselling, treatment and care for adolescents living with HIV. Geneva: WHO2013.

[3] World-Bank. Ethiopia – Prevalence of HIV as a share of population aged 15–49. World Data Atlas.. 2021.

[4] UNICEF. 2021 HIV and AIDS in sub-Saharan Africa snapshot: Pregnant women, children, and adolescents.. 2021.

[5] Bongaarts J. FAO, IFAD, UNICEF, WFP and WHO The State of Food Security and Nutrition in the World 2020. Transforming food systems for affordable healthy diets FAO, 2020, 320 p. New York: Wiley Subscription Services, Inc; 2021. p. 558-.

[6] Norris SA, Frongillo EA, Black MM, Dong Y, Fall C, Lampl M, et al. Nutrition in adolescent growth and development. The Lancet. 2022;399(10320):172–84.

[7] Abera M, Hardy-Johnson P, Abdissa A, Workicho A, Ali R, Weller S, et al. Social, economic and cultural influences on adolescent nutrition and physical activity in Jimma, Ethiopia: perspectives from adolescents and their caregivers. Public health nutrition. 2021;24(16):5218-26.

[8] FANTA. Defining Nutrition Assessment, Counseling, and Support (NACS). Technical Note No. 13. Washington, DC: FHI 360/FANTA. 2012 [Available from: https://www.fantaproject.org/focus-areas/infectious-diseases/nacs-technical-note.

[9] WHO. Nutrient requirements for people living with HIV/AIDS : report of a technical consultation, 13-15 May 2003, Geneva. Geneva: World Health Organization; 2004.

[10] National guideline on adolescent, maternal, infant and young child nutrition. Addis Ababa, Ethiopia: Federal Democratic Republic of Ethiopia, Ministry of Health [Internet]. 2016. Available from: https://www.dataverse.nipn.ephi.gov.et/bitstream/handle/123456789/1034/AMIYCN%20Guideline%202016.pdf.

[11] Endris B, Fenta E, Getnet Y, Spigt M, Dinant GJ, Gebreyesus S. Barriers and facilitators to the implementation of nutrition interventions at primary health care units of Ethiopia: A consolidated framework for implementation research. Maternal & Child Nutrition. 2022;19.

[12] Elm Ev, Altman DG, Egger M, Pocock SJ, Gøtzsche PC, Vandenbroucke JP. Strengthening the reporting of observational studies in epidemiology (STROBE) statement: guidelines for reporting observational studies. BMJ. 2007;335(7624):806-8.

[13] Lachat C, Hawwash D, Ocké MC, Berg C, Forsum E, Hörnell A, et al. Strengthening the Reporting of Observational Studies in Epidemiology-Nutritional Epidemiology (STROBE-nut): An Extension of the STROBE Statement. PLoS Med. 2016;13(6):e1002036.

[14] Vandenbroucke JP, von Elm E, Altman DG, Gøtzsche PC, Mulrow CD, Pocock SJ, et al. Strengthening the Reporting of Observational Studies in Epidemiology (STROBE): Explanation and elaboration. International Journal of Surgery. 2014;12(12):1500–24.

[15] Shiferaw H, Gebremedhin S. Undernutrition Among HIV-Positive Adolescents on Antiretroviral Therapy in Southern Ethiopia. Adolesc Health Med Ther. 2020;11:101–11.

[16] Kennedy-Hagan K. R. Gibson Principles of Nutritional Assessment 2 nd ed. 2005 From Oxford University Press, Inc, 198 Madison Ave, New York, NY 10016, (212) 726-6000, hardcover, 908 pp, $95.00, ISBN 0-19-517169-1. Elsevier Inc; 2006. p. 331–2.

[17] Aldobali M, Pal K, editors. Bioelectrical Impedance Analysis for Evaluation of Body Composition: A Review2021 2021: IEEE.

[18] Bohannon R. Grip Strength: A Summary of Studies Comparing Dominant and Nondominant Limb Measurements. Perceptual and motor skills. 2003;96:728–30.

[19] Pham MD, Nguyen HV, Anderson D, Crowe S, Luchters S. Viral load monitoring for people living with HIV in the era of test and treat: progress made and challenges ahead – a systematic review. BMC Public Health. 2022;22(1):1203.

[20] WHO. Considerations for developing a monitoring and evaluation framework for viral load testing. Geneva: World Health Organization; 2019 (WHO/CDS/HIV/19.5). Licence: CC BY-NC-SA 3.0 IGO. 2019 [

[21] Bhaskaran K, Hamouda O, Sannes M, Boufassa F, Johnson AM, Lambert PC, et al. Changes in the risk of death after HIV seroconversion compared with mortality in the general population. JAMA. 2008;300(1):51–9.

[22] Kroenke K, Spitzer RL, Williams JB. The PHQ-9: validity of a brief depression severity measure. J Gen Intern Med. 2001;16(9):606–13.

[23] Swinson RP. The GAD-7 scale was accurate for diagnosing generalised anxiety disorder. Evid Based Med. 2006;11(6):184.

[24] AUDIT: The Alcohol Use Disorders Identification Test: Guidelines for Use in Primary Care (2nd ed.). Geneva: World Health Organization [Internet]. 2001. Available from: https://apps.who.int/iris/bitstream/10665/67205/1/WHO_MSD_MSB_01.6a.pdf.

[25] Higgins-Biddle JC, Babor TF. A review of the Alcohol Use Disorders Identification Test (AUDIT), AUDIT-C, and USAUDIT for screening in the United States: Past issues and future directions. Am J Drug Alcohol Abuse. 2018;44(6):578–86.

[26] Moussas G, Dadouti G, Douzenis A, Poulis E, Tzelembis A, Bratis D, et al. The Alcohol Use Disorders Identification Test (AUDIT): reliability and validity of the Greek version. Ann Gen Psychiatry. 2009;8:11.

[27] McHugh ML. Interrater reliability: the kappa statistic. Biochem Med (Zagreb). 2012;22(3):276–82.

[28] Sidani S, Guruge S, Miranda J, Ford-Gilboe M, Varcoe C. Cultural adaptation and translation of measures: an integrated method. Research in nursing & health. 2010;33 2:133–43.

[29] Yusoff MSB. ABC of content validation and content validity index calculation. Resource. 2019;11(2):49–54.

[30] Polit DF, Beck CT, Owen SV. Is the CVI an acceptable indicator of content validity? Appraisal and recommendations. Res Nurs Health. 2007;30(4):459–67.

[31] Adib Y, Fathiazar E, Alizadeh M, Dehghani G. Development and Validation of Instruments for Evaluation of the Clinical Medicine Curriculum in Terms of Social Accountability. Journal of Medical Education Development. 2018;11:1–12.

[32] Iheme G, Uzokwe C, Ezenwa H, Nwamadi C, Okonkwo E, Matthew S. Application of WHO 2007 growth reference in assessing the anthropometric status of Nigerian adolescents; A systematic review and meta analysis. Human Nutrition & Metabolism. 2022;28:200144.

[33] FANTA-III. BMI and BMI-for-Age Look-Up Tables for Children and Adolescents 5–18 Years of Age and BMI Look-Up Tables for Non-Pregnant, Non-Lactating Adults ≥ 19 Years of Age 2013

[34] WHO. Growth reference data for 5-19 years 2007

[35] WHO. WHO Child Growth Standards based on length/height, weight and age. Acta Paediatr Suppl. 2006;450:76–85.

[36] Kristen C, Lesley O. Guide to anthropometry: a practical tool for program planners, managers, and implementers. Food and Nutrition Technical Assistance III Project (FANTA)/FHI. 2018;360:2020–03.

[37] De Onis M, Onyango AW, Borghi E, Siyam A, Nishida C, Siekmann J. Development of a WHO growth reference for school-aged children and adolescents. Bulletin of the World Health Organization. 2007;85(9):660–7.

[38] Christian P, Smith ER. Adolescent Undernutrition: Global Burden, Physiology, and Nutritional Risks. Ann Nutr Metab. 2018;72(4):316–28.

[39] Francis L, Wanyenze R, Matovu J, Chimulwa T, Orach C. Nutritional Status of HIV-infected Adolescents Enrolled into an HIV-care Program in Urban and Rural Uganda: A Cross-sectional Study. Journal of Nutrition and Health. 2015;3:29–34.

[40] Fagbamigbe AF, Adebowale AS, Ajayi I. An assessment of the nutritional status of ART receiving HIV-orphaned and vulnerable children in South-West Nigeria. Heliyon. 2019;5(12):e02925.

[41] Dave DA, Provia A, Nakiddu N, Sodawasser E, Harper K, Ssenkusu J, et al. Nutritional status and its associated factors among HIV adolescents on second line regimen at Pediatric Infectious Diseases Clinic in Uganda. Journal of HIV/AIDS & Social Services. 2022;21(1):63–75.

[42] WHO. Nutrient requirements for people living with HIV/AIDS : report of a technical consultation, World Health Organization, Geneva, 13–15 May 2003. 2003.

[43] Fathima AS, Madhu M, Udaya Kumar V, Dhingra S, Kumar N, Singh S, et al. Nutritional Aspects of People Living with HIV (PLHIV) Amidst COVID-19 Pandemic: an Insight. Curr Pharmacol Rep. 2022;8(5):350–64.

[44] Saavedra JM, Prentice AM. Nutrition in school-age children: a rationale for revisiting priorities. Nutr Rev. 2023;81(7):823–43.

[45] de Pee S, Semba RD. Role of nutrition in HIV infection: review of evidence for more effective programming in resource-limited settings. Food Nutr Bull. 2010;31(4):S313–44.

[46] Semba RD, Darnton-Hill I, de Pee S. Addressing tuberculosis in the context of malnutrition and HIV coinfection. Food Nutr Bull. 2010;31(4):S345–64.

[47] Melku M, Enawgaw B, Ayana S, Anlay DZ, Kebede A, Haile A, et al. Magnitude of anemia and undernutrition among HIV-infected children who took HAART: a retrospective follow-up study. Am J Blood Res. 2020;10(5):198–209.

[48] Saunders J, Brian A, Wright M, Stroud M. Malnutrition and nutrition support in patients with liver disease. Frontline Gastroenterol. 2010;1(2):105–11.

[49] Martín-Cañavate R, Sonego M, Sagrado MJ, Escobar G, Rivas E, Ayala S, et al. Dietary patterns and nutritional status of HIV-infected children and adolescents in El Salvador: A cross-sectional study. PLoS One. 2018;13(5):e0196380.

[50] Holmes CJ, Racette SB. The Utility of Body Composition Assessment in Nutrition and Clinical Practice: An Overview of Current Methodology. Nutrients. 2021;13(8).

[51] Bain R, Cronk R, Hossain R, Bonjour S, Onda K, Wright J, et al. Global assessment of exposure to faecal contamination through drinking water based on a systematic review. Tropical medicine & international health. 2014;19(8):917–27.

[52] Hutton G, Haller L, Bartram J. Global cost-benefit analysis of water supply and sanitation interventions. Journal of water and health. 2007;5(4):481–502.

[53] Fan Y, Yao Q, Liu Y, Jia T, Zhang J, Jiang E. Underlying Causes and Co-existence of Malnutrition and Infections: An Exceedingly Common Death Risk in Cancer. Frontiers in Nutrition. 2022;9.

[54] Michel M, Labenz C, Armandi A, Kaps L, Kremer WM, Galle PR, et al. Metabolic dysfunction-associated fatty liver disease in people living with HIV. Scientific Reports. 2023;13(1):9158.

[55] Sawyer SM, Azzopardi PS, Wickremarathne D, Patton GC. The age of adolescence. The Lancet Child & Adolescent Health. 2018;2(3):223–8.

[56] Suto M, Miyazaki C, Yanagawa Y, Takehara K, Kato T, Gai R, et al. Overview of Evidence Concerning School-Based Interventions for Improving the Health of School-Aged Children and Adolescents. The Journal of school health. 2021;91(6):499–517.

[57] Xu T, Tomokawa S, Gregorio ER, Jr., Mannava P, Nagai M, Sobel H. School-based interventions to promote adolescent health: A systematic review in low- and middle-income countries of WHO Western Pacific Region. PLoS One. 2020;15(3):e0230046.

